# Acquisition of extended-spectrum beta-lactamase-producing Enterobacteriaceae (ESBL-PE) carriage after exposure to systemic antimicrobials during travel: systematic review and meta-analysis

**DOI:** 10.1101/19007781

**Authors:** Terence C. Wuerz, Sameer S. Kassim, Katherine E. Atkins

## Abstract

**Background:** International travel is an important risk factor for colonization with extended-spectrum beta-lactamase-producing Enterobacteriaceae (ESBL-PE). Antimicrobial use during travel likely amplifies this risk, yet to what extent, and whether it varies by antimicrobial class, has not been established.

**Methods:** We conducted a systematic review that included prospective cohorts reporting both receipt of systemic antimicrobials and acquired ESBL-PE isolated from stool or rectum during international travel. We performed a random effects meta-analysis to estimate odds of acquiring ESBL-PE due to antimicrobials during travel, overall and by antimicrobial class.

**Results:** Fifteen studies were included. The study population was mainly female travellers from high income countries recruited primarily from travel clinics. Participants travelled most frequently to Asia and Africa with 10% reporting antimicrobial use during travel. The combined odds ratio (OR) for ESBL-PE acquisition during travel was 2.37 for antimicrobial use overall (95% confidence interval [CI], 1.69 to 3.33), but there was substantial heterogeneity between studies. Fluoroquinolones were the antibiotic class associated with the highest combined OR of ESBL-PE acquisition, compared to no antimicrobial use (OR 4.68, 95% CI, 2.34 to 9.37).

**Conclusions:** The risk of ESBL-PE colonization during travel is increased substantially with exposure to antimicrobials, especially fluoroquinolones. While a small proportion of colonized individuals will develop a resistant infection, there remains the potential for onward spread among returning travellers. Public health efforts to decrease inappropriate antimicrobial usage during travel are warranted.

**Research in context:** *Evidence before this study:* Antimicrobial resistance (AMR) among bacteria that commonly cause human infection is of increasing public health concern. International travel has recently been associated with colonization with Extended-Spectrum Beta-Lactamase Producing-Enterobacteriaceae (ESBL-PE), increasing the spread of drug resistance among these important pathogens. We searched Pubmed, Embase, MEDLINE, Web of Science, SCOPUS, and the Cochrane Library for prospective cohort studies published between January 2000 and June 2018, reporting on acquisition of ESBL-PE among travellers, which reported on antimicrobial use during travel. 15 studies were included, which were at moderate risk of bias. The pooled odds ratio for acquisition of ESBL-PE during travel was 2.37 among antimicrobial users, compared to non-users (95% CI, 1.69 to 3.33). The magnitude of this association was stronger among travellers reporting fluoroquinolone use (OR 4.68, 95% CI 2.34 to 9.37).

*Added value of this study:* This is the first study to quantify the association between antimicrobial use during travel, overall and by specific antimicrobial class, with ESBL-PE acquisition across broad populations of travellers and destination countries.

*Implications of all the available evidence:* Further study into the mechanisms by which antimicrobials, such as fluoroquinolones, contribute to AMR may identify protective measures. Meanwhile, antimicrobial use during travel for prevention or treatment of mild-to-moderate traveller’s diarrhea should not be recommended routinely. Where indicated, alternatives to fluoroquinolone antimicrobials should be considered.

## INTRODUCTION

In 2014 the World Health Organization declared that a “post-antibiotic era” is within sight if urgent action was not taken^1^. At the forefront of this growing public health threat is drug resistance in Enterobacteriaceae, a family of gram-negative bacteria which make up a large part of normal human gut flora. Extended-spectrum beta-lactamase (ESBL) enzymes are an important cause of increasing bacterial resistance globally^2,3^ with ESBL-producing Enterobacteriaceae (ESBL-PE) colonizing humans either de novo or through fecal-oral transmission^4^. ESBL enzymes render all penicillin, monobactam and expanded-spectrum cephalosporin antimicrobials^2,5^ ineffective. Carriers of are usually asymptomatic^4^, but are at risk to develop clinical infection due to these organisms, resulting in increased cost, morbidity, and mortality compared to infections due to drug-susceptible Enterobacteriaceae^4,6,7^.

The prevalence of ESBL-PE varies worldwide and by epidemiologic setting^1,3,4^. For example, the frequency of ESBL-PE is relatively low in North America and Europe^1^, and comparatively much higher in many lower income countries, especially in South Asia^4^. Accordingly, observational studies have established a strong association between asymptomatic ESBL-PE acquisition and international travel^8,9,18–23,10–17^. The risk of acquiring carriage with ESBL-PE is over 20% with any international travel, and it is higher with travel to areas of particularly high prevalence such as South Asia^23^. Although most healthy returning travellers colonized with ESBL-PE will not develop infection with these bacteria, some do^24,25^, and an estimated 12% will transmit these bacteria onwards to other household members^9^.

Antimicrobial use has been inconsistently reported as an independent risk factor for ESBL-PE acquisition among international travellers^23^. A previous review noted that 4 of the 11 included studies found a significant association between antimicrobial use during travel and faecal ESBL-PE acquisition. However, this study did not quantify this association, nor did it examine the role that different antimicrobial classes play in the risk of ESBL-PE acquisition.

We conducted a systematic review and meta-analysis to determine the extent to which receipt of antimicrobials during travel, as well as which classes of antimicrobials, increase the odds of faecal carriage with ESBL-PE compared to those that do not receive antimicrobials.

## METHODS

We included only prospective cohort studies related to travel across an international boundary reporting screening for faecal Enterobacteriaceae carriage (in asymptomatic participants) both prior to and after travel, in our review. Studies must have presented phenotypic antimicrobial susceptibility data (or molecular equivalent) which is adequate to ascertain ESBL-PE presence or absence, and were excluded if ESBL-PE acquisition status by presence or absence of receipt of systemic antimicrobials taken during travel was not reported.

We searched Embase, MEDLINE, Web of Science, SCOPUS, Cochrane Library, and PubMed for studies published in peer-reviewed journals from January 2000 to June 2018 (Box 1). The reference lists, and citations of included studies, were inspected to identify further potentially eligible studies. Citations were entered and housed in Covidence^™^, and abstracts were screened for eligibility by 2 independent reviewers (TW, SK). Studies selected for full text review were read in full to ascertain whether inclusion criteria were met and selected for data extraction as appropriate. Conflicts were resolved by consensus by 2 authors (TW, SK). Each included study was assessed using tools for assessing bias in observational studies as recommended in the STROBE statement^26^ and modified to suit the study design.

Data were extracted by a single author (TW) from each included article and exported to Stata 13 for analysis. Data gathered included details on study population and travel characteristics, potential confounders, antimicrobials taken during travel, method of determining ESBL-PE pre- and post-travel, the prevalence of ESBL-PE pre- and post-travel, and study quality metrics. Where possible, percentages not presented in the original articles were calculated from available data. Odds ratios for acquisition of ESBL-PE, by presence or absence of antimicrobial exposure during travel, with 95% confidence intervals of all included studies was displayed as a Forrest plot. Data were visually inspected for heterogeneity of results and Cochrane’s Q and I^2^ were calculated. As there was substantial evidence of heterogeneity, a random effects model of meta-analysis, representing the average odds ratio of ESBL-PE due to antimicrobial exposure during travel, was built. For the secondary analysis, a random effects meta-analysis was undertaken of rates of ESBL-PE acquisition by antimicrobial class (beta-lactam, fluoroquinolone, macrolide or tetracycline) received during travel, compared to no antimicrobial receipt (the baseline group).

All antimicrobials including doxycycline were included in the primary analysis, however, doxycycline was excluded from this analysis in 2 studies, as it was counted as an antimalarial drug. We therefore performed a sensitivity analysis by comparing the random effects meta-analysis which excluded doxycycline as an antimicrobial to the primary analysis which included. For all studies, we plotted effect size (log OR for ESBL-PE acquisition during travel related antimicrobial exposure) against log standard error of each study to create a funnel plot. The quality of the studies was assessed using the modified STROBE^26^ tool.

Studies were excluded from the secondary analysis if no information was available on proportion of ESBL-PE acquisition stratified by antimicrobial class. Authors of studies missing crucial data were contacted to obtain it.

## RESULTS

The search strategy identified 5323 journal articles in published peer-reviewed journals (Figure 1). 1893 duplicates were identified and removed, leaving 3430 study titles and abstracts, of which 3372 were excluded on screening. 58 published manuscripts were reviewed in full and 15 studies were included for the systematic review.

**Figure 1:**
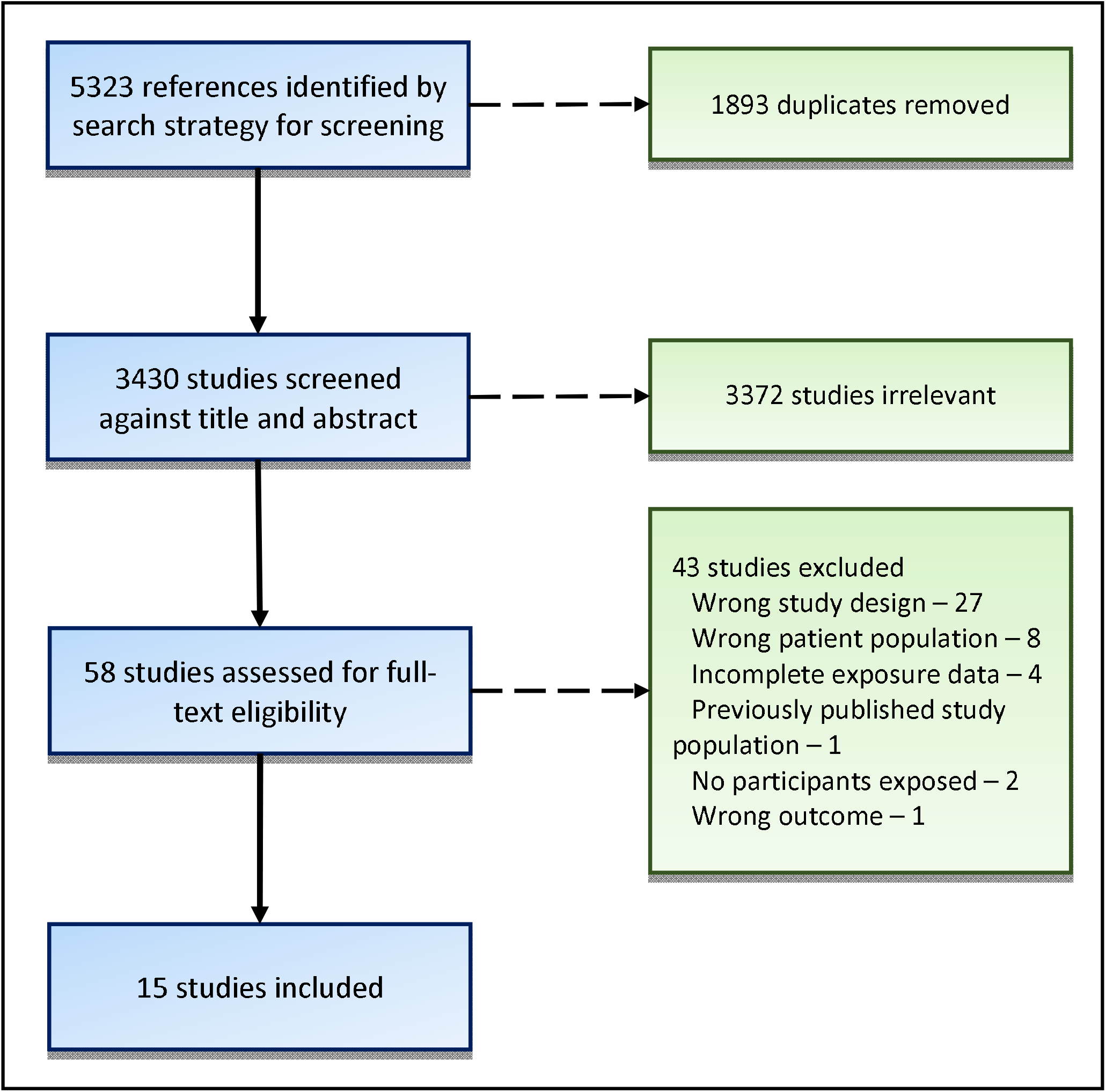
Flowchart depicting studies screened, reviewed and finally included for systematic review.

The 15 included prospective cohort studies were published from 2010 to 2017, and enrolled travellers from 2007 to 2015^8,9,18–22,10–17^. The studies included a median of 205 participants (range, 58 to 1965) and in all but 2 instances were conducted in Northern or Western Europe (Table 1a). The median age of participants varied substantially, but in all studies, fewer than 50% were male. 13 studies used a phenotypic approach for determination of ESBL-PE status (with polymerase chain reaction [PCR] for confirmation) whereas 2 studies adopted a molecular approach, using PCR to identify the gene encoding ESBL production within Enterobacteriaceae, bla_CTX-M_, without culture^14,22^.

**Table 1a:**
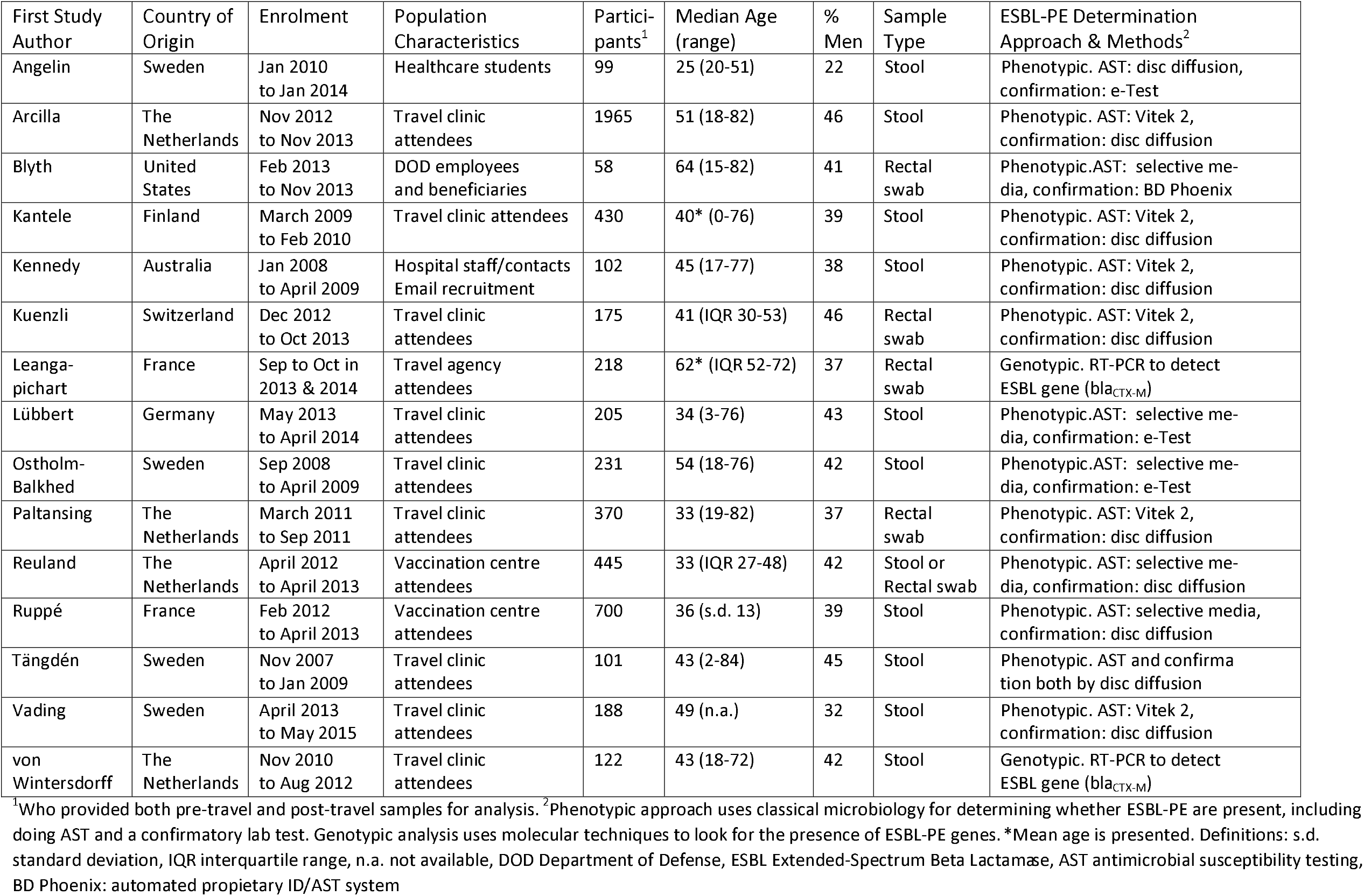
Key methodology and baseline characteristics for studies included for systematic review of the risk of acquiring Extended-Spectrum Producing-Enterobacteriaceae after exposure to antimicrobials during travel.

**Table 1b:** Characteristics of travel for studies included for systematic review of the risk of acquiring Extended-Spectrum Producing-Enterobacteriaceae after exposure to antimicrobials during travel.

| First Study Author | Top 3 Regions Travelled to<sup>2</sup><br>(Percentage of Participants<sup>1</sup>) | Travel Time Median in Days<br>(range) | Percentage of all travellers who: |  |  | Top 2 Classes of Antimicrobials Used (% Total) |
| --- | --- | --- | --- | --- | --- | --- |
|  |  |  | Had Diarrhea | Used Any Abtx | Acquired ESBL-PE |  |
| Angelin | South Asia(40), SSA (39), Asia(8) | 45<br>(13-365) | 69 | 20 | 36 | Beta-lactams (30),<br>FQs (20) |
| Arcilla | Asia (41), SSA (21), South/Central America (18) | 20<br>(15-25†) | 39 | 7 | 34 | FQs (31), Beta-lactams (20) |
| Blyth | South/Central America (33), Asia & South Asia (27), Africa (27) | 12*<br>(6-105) | 12 | 10 | 9 | Tetracyclines (50), FQs (33%) |
| Kantele | SSA (45), Asia (25), South Asia (14) | 19<br>(4-133) | 67 | 15 | 21 | FQs (70),<br>Macrolides (14) |
| Kennedy | Asia (49), Europe (16), South Asia (11) | 21<br>(9-135) | 37 | 27 | 49 | Tetracyclines (54), FQ (7) |
| Kuenzli | South Asia (100) | 18<br>(5-35) | 37 | 4 | 69 | FQs (43), Beta-lactams (29) |
| Leanga-pichart | Middle East (100) | 28<br>(24-32) | 19 | 49 | 37 | Beta-lactams (72)<br>Macrolides (29) |
| Lübbert | SSA (31), Asia (30), South/Central America (27) | 21<br>(3-218) | 38 | 13 | 30 | Tetracyclines (38), FQs (17) |
| Ostholm-Balkhed | Africa (34), Asia (26)<br>South & Central America (13) | 16<br>(4-119) | 42 | 9 | 30 | NR |
| Paltansing | Asia (43), South Asia (25), SSA (25) | 21<br>(6-90) | 38 | 6 | 33 | Beta-lactams (32), FQs (21) |
| Reuland | Asia & South Asia (56), Africa (22), South/Central America (18) | NR | 46 | 5 | 23 | FQs (27), Beta-lactams (18) |
| Ruppé | Asia & South Asia (34), SSA (34), South/Central America (31) | 20<br>(15-30) | 40 | 10 | 51 | Beta-lactams (42),<br>FQs (22) |
| Tängdén | Asia (30), SSA (24), Europe (15) | 14<br>(1-26) | 30 | 10 | 24 | Beta-lactams (50),<br>FQs (30) |
| Vading | Asia (39), South Asia (35), Northern Africa (14) | 14*<br>(8-20†) | 29 | 9 | 24 | FQs (33), Beta-lactams (20) |
| von Wintersdorff | Africa (27), South Asia (25), Asia (23) | 21<br>(5-240) | 37 | 14 | 31 | NR |

<sup>1</sup>Who provided both pre-travel and post-travel samples for analysis. <sup>2</sup>Unless otherwise indicated, South Asia (including India, Pakistan, Nepal, Sri Lanka, and Bangladesh) included separately from the rest of Asia, due to higher rates of ESBL-
PE. \*Median is presented. †Interquartile range is presented. Definitions: NR not reported, ESBL-PE Extended-spectrum beta-lactamase-producing Enterobacteriaceae, SSA Sub-Saharan Africa, FQ fluoroquinolone

**Table 1c:** Rates oxtended-spectrum beta-lactamase acquisition by antimicrobial exposure during travel, with crude odds ratio and 95% confidence interval, for included studies.

| First Study<br>Author | # participants<sup>1</sup><br>who acquired<br>ESBL-PE (% all<br>participants) | Number exposed to<br>antibiotics who<br>acquired ESBL-PE<sup>2</sup><br>(% of those exposed) | Number not exposed<br>to antibiotics who<br>acquired ESBL-PE<sup>2</sup> (%<br>of those not exposed) | Odds ratio for<br>acquisition of ESBL-PE by antibiotic<br>exposure (95% CI) |
| --- | --- | --- | --- | --- |
| Angelin | 35 (36) | 20 (25) | 30 (38) | 0.53 (0.18, 1.62) |
| Arcilla | 633 (34) | 132 (55) | 553 (33) | 2.56 (1.79, 3.66) |
| Blyth | 5 (9) | 6 (33) | 3 (6) | 8.50 (1.09, 66.6) |
| Kantele | 90 (21) | 66 (42) | 62 (17) | 3.53 (2.02, 6.18) |
| Kennedy | 50 (49) | 28 (68) | 31 (42) | 2.93 (1.17, 7.33) |
| Kuenzli | 118 (69) | 7 (71) | 101 (69) | 1.14 (0.21, 6.09) |
| Leangapichart | 73 (37) | 107 (36) | 35 (40) | 0.83 (0.47, 1.49) |
| Lübbert | 58 (30) | 24 (38) | 15 (9) | 6.08 (2.28, 16.23) |
| Ostholm-Balkhed | 68 (30) | 20 (25) | 15 (7) | 4.24 (1.36, 13.28) |
| Paltansing | 113 (33) | 19 (47) | 104 (33) | 1.86 (0.73, 4.72) |
| Reuland | 95 (23) | 22 (50) | 87 (22) | 3.46 (1.45, 8.25) |
| Ruppé | 292 (51) | 59 (73) | 249 (48) | 2.87 (1.58, 5.23) |
| Tängdén | 24 (24) | 10 (30) | 21 (23) | 1.41 (0.33, 5.93) |
| Vading | 56 (24) | 15 (67) | 46 (29) | 4.91 (1.59, 15.16) |
| Von Wintersdorff | 38 (31) | 15 (40) | 32 (30) | 1.56 (0.51, 4.75) |

<sup>1</sup>Who provided both pre-travel and post-travel samples for analysis.
<sup>2</sup>Excluding participants with missing antibiotic exposure information.

Asia, including South Asia region to which travel carries the highest risk of ESBL-PE acquisition, was the most common region travelled to, followed by Africa (Table 1b). Median travel time ranged from 14-21 days in most studies, and the median percentage of travellers reporting diarrhea or gastroenteritis was 38% (range: 12 to 69%). We found a median of reported systemic antimicrobial usage of 10% (range: 4-49%). The most common indications (where reported) were lower respiratory tract infection and traveller’s diarrhea. Of the 11 studies reporting antimicrobial class used, beta-lactam and fluoroquinolone antimicrobials were the most common classes. The percentage of travellers who acquired ESBL-PE carriage during travel also varied significantly by study, from 9% of to 69% (median: 31%).

All studies used reliable laboratory phenotypic or genotypic methodology; however, all enrolment was from non-random samples of travellers. All but 2 studies^17,18^ did not exclude co-travellers (groups of travellers) from enrolment. The exposure of interest, antimicrobial use during travel, was ascertained via post-travel questionnaire all studies. While important potential confounding was reported in all studies, duration of antimicrobial usage was not reported by any. 4 studies^10,17,22,27^ provided no information about losses to follow-up; for the remainder, the median number lost to follow-up was low (7.5%).

The point estimates of the odds ratio (OR) for ESBL-PE acquisition by any or no systemic antimicrobial exposure varied between 0.53 to 8.05 for individual studies (Table 1c).

We included all studies in a random effects meta-analysis (Figure 2). Our primary analysis found that the combined OR for effect of all antimicrobials on ESBL-PE acquisition was 2.37 (95% CI 1.69 to 3.33, p < 0.01, I^2^=57%, which indicates strong evidence for heterogeneity). A sensitivity analysis excluding doxycycline (all but 2 studies^15,16^), yielded very similar results (OR 2.48, 95% CI 1.76 to 3.50, p<0.01, I^2^ 54%). Visual inspection of the funnel plot (Figure 3) found no strong evidence of bias across the studies.

**Figure 2:**
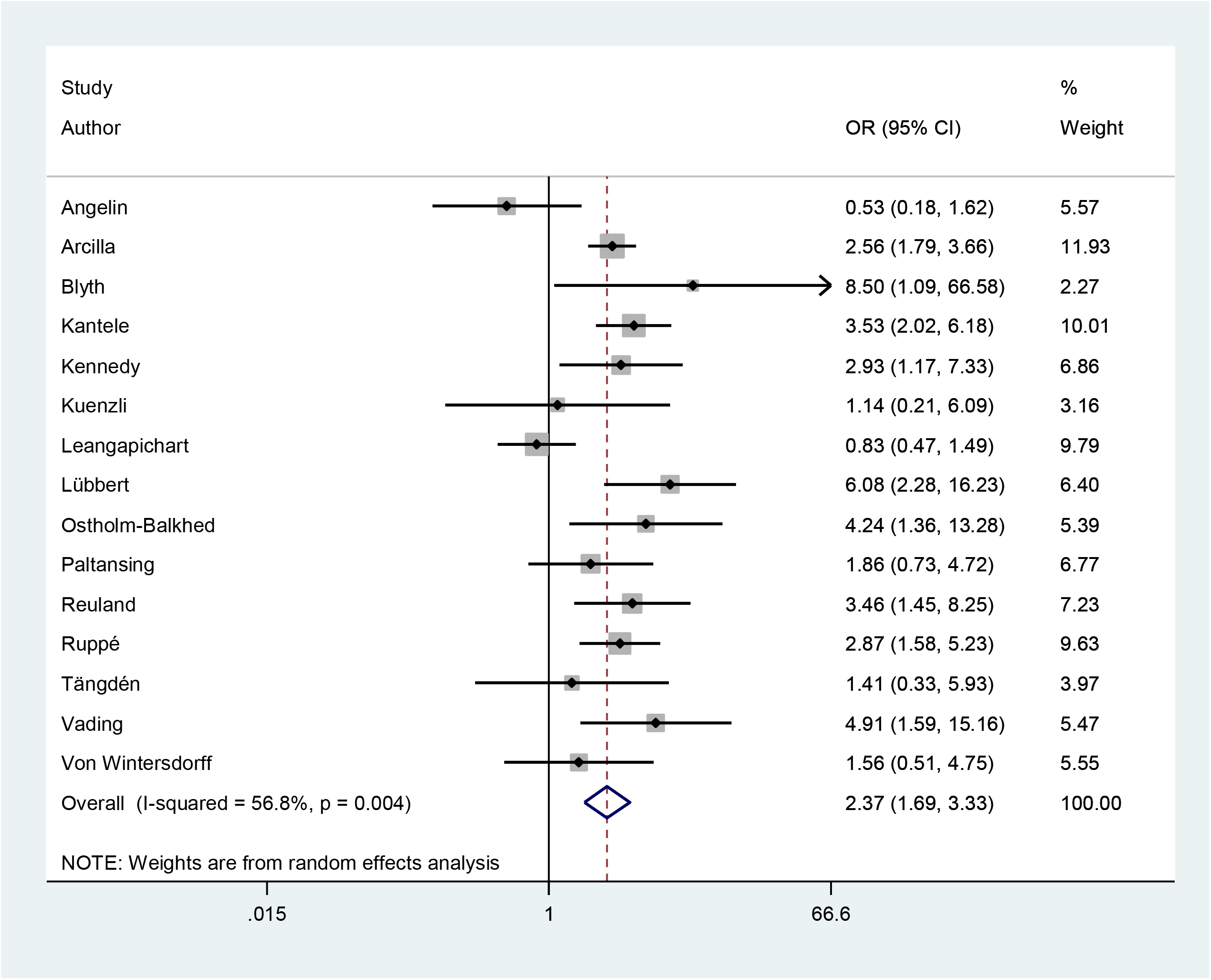
Forest plot of random effects model, for odds ratio (OR) of extended-spectrum beta-lactamase-producing Enterobacteriaceae (ESBL-PE) among participants exposed to, compared to those not exposed to, antimicrobials during travel. Definitions: OR odds ratio, CI confidence intervals.

**Figure 3:**
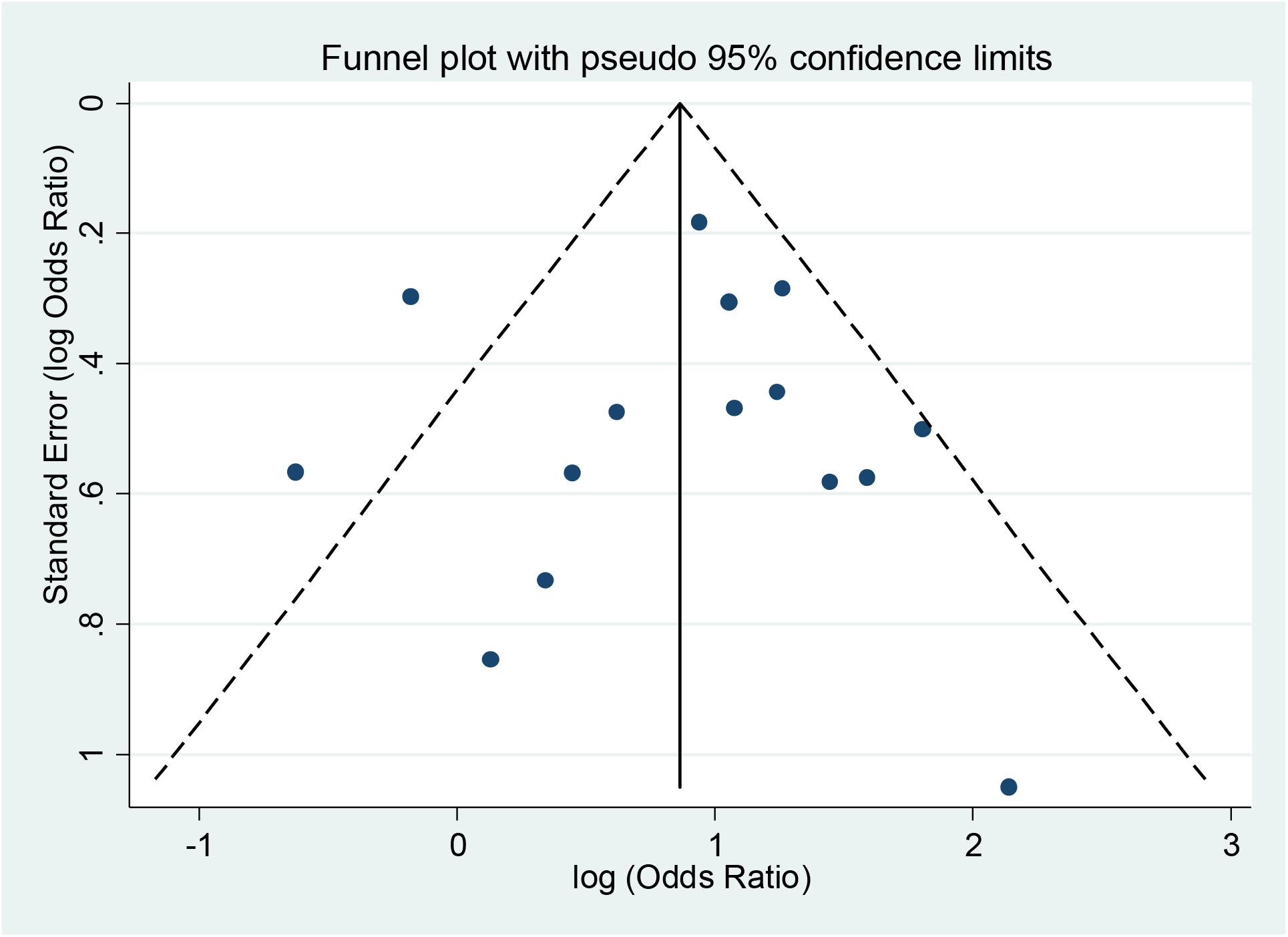
Funnel plot of included studies with the effect size (Odds Ratio (OR) for Extended-Spectrum Beta-Lactamase-Producing Enterobacteriaceae (ESBL-PE) acquisition during travel due to antimicrobial exposure) plotted against standard error*. *Each point on the figure represents an included study with its OR and sample size. Interposed triangle is centred on a fixed effects meta-analysis summary log OR. Pseudo 95% confidence limits are the dashed lines which represent 1.96 standard errors extending to either side of the summary log OR.

Excluding the 4 studies that had no information on ESBL-PE acquisition by class of antimicrobial acquisition by class of antimicrobial received^11,16,21,22^, we found fluoroquinolone exposure was associated with a combined OR of 4.68 compared to no antibiotics (95% CI, 2.34 to 9.37), tetracyclines were associated a combined OR of 1.68 (95% CI, 1.03 to 2.72) (Table 2). On average, there was no evidence of a combined increased odds of ESBL-PE acquisition by exposure to beta-lactams or macrolides.

**Table 2:**
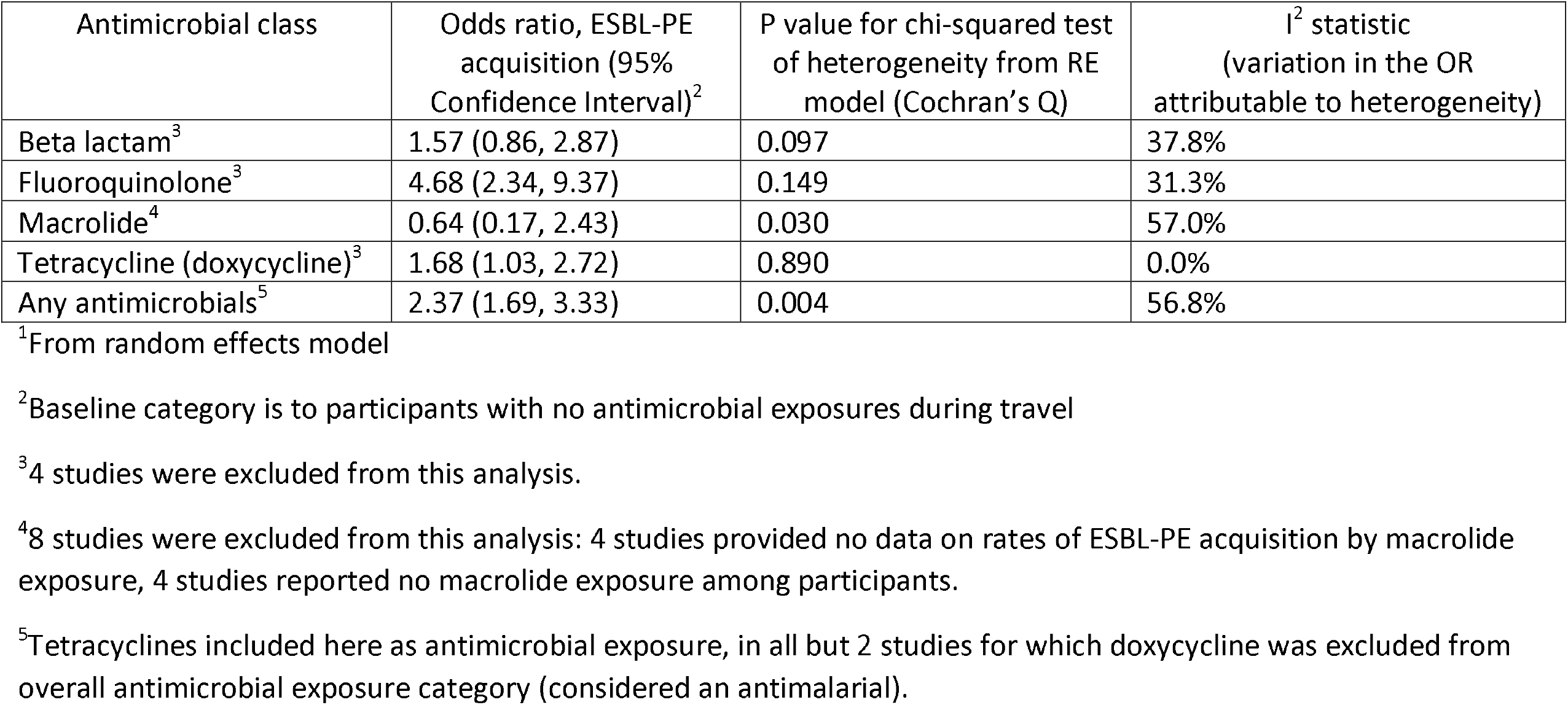
Summary measure of effect, with 95% confidence intervals and measures of heterogeneity, for odds of acquiring extended-spectrum beta-lactamase-producing Enterobacteriaceae (ESBL-PE) during travel, with exposure to specific classes of antimicrobials during travel across all included studies^1^

## DISCUSSION

International travel has been previously identified as an important risk factor for both colonization and infection with ESBL-PE^23–25^, and likely plays an important role in the spread of ESBL-PE within high income countries. However, the risk has not been systematically quantified. Our systematic review and meta-analysis found that systemic antimicrobial use during travel was associated with an odds ratio of 2.37 for acquiring ESBL-PE. Given a baseline risk of ESBL-PE acquisition of 20% or higher among international travellers^9,23^, and antimicrobial use in an average of 10% of such travellers, the burden of this additional risk is substantial. Surprisingly, our analysis found that fluoroquinolones, which are not of the beta-lactam family of antimicrobials, were associated with the highest risk of ESBL-PE acquisition, with a combined OR of 4.68.

While antimicrobial use is widely recognized to be a major driver of AMR, quantifying its impact can be challenging. Ecologic data comparing antimicrobial consumption in the community and hospital has been positively correlated to AMR (from hospital bacterial isolates) over time and across countries^28,29^. A previous systematic review^23^ examining the risk of ESBL-PE acquisition in travellers including 11 of the 15 studies included here described an association between antimicrobials taken during travel and ESBL-PE acquisition in 4 studies. Unlike the previous study, we use a meta-analysis to estimate the degree to which antimicrobial usage during travel increases the odds of ESBL-PE acquisition across populations of international travellers from high income countries and providing insight into the relatively high odds attributable specifically to travellers using fluoroquinolone antimicrobials.

There exist several pathways through which antimicrobial use might cause an increase in acquisition in ESBL-PE during travel. First, selection pressure due to antimicrobial effect on more drug-sensitive strains of Enterobacteriaceae can select for increased resistance within an individual. This selection could be due to antimicrobials taken in response to disease causing Enterobacteriaceae or, substantially more likely, due to the by-stander effect whereby non-invasive bacteria are exposed to antimicrobials^30^. This pathway would explain why beta-lactams, but not fluroquinolones can be causatively associated with an increased acquisition of ESBL-PE. An alternate pathway involves the ability of the healthy gut microbiota to prevent expansion of potential pathogens including resistant bacteria such as ESBL-PE, a phenomenon termed ‘colonization resistance’^31^. Disruption of microbial composition due to exposure to antimicrobials decreases colonization resistance^32^, reducing the normally protective capabilities of an intestine with fully diverse and intact microbial composition. Proposed mechanisms for the normal flora-mediated resistance to colonization include direct microbial competition for nutrients, production of bacteriocin peptides which inhibit the growth of specific types of bacteria, and more complex indirect mechanisms involving interaction between bacterial communities to help maintain host immune responses^32,33^. Fluoroquinolones have a relatively large impact on the composition of normal gut microbiota composition due to their broad spectrum of activity and high local concentrations achieved^34^, which may in part explain their large effect on ESBL-PE acquisition seen in this study.

An estimated 30% of outpatient antimicrobial prescriptions in the US are considered inappropriate^35^. It is likely that inappropriate antimicrobial use is even higher in lower income countries, where most consumption is attributable to over-the-counter use. Moreover, travellers from high income countries to tropical settings have historically been counselled to bring antimicrobials, including commonly fluoroquinolones, as preventative or abortive treatment of diarrhea^36,37^. While more recent guidelines have scaled back the uniform recommendation of traveller antimicrobial use, partly in response to increased recognition of the risk of faecal drug resistant carriage, they still suggest use of antimicrobial chemoprophylaxis in some higher-risk travellers, and antimicrobial therapy for all cases of moderate-to-severe traveller’s diarrhea^36^. Loperamide offers an alternative for the management of mild-to-moderate traveller’s diarrhea which appears to be as effective as antimicrobials but does not increase the risk of colonization with ESBL-PE^38^. This study supports calls^39^ to avoid unnecessary antimicrobial consumption during travel, including for prevention or treatment of mild-to-moderate severity traveller’s diarrhea in healthy travellers. When they cannot be avoided altogether, our results suggest using alternative antimicrobials to fluoroquinolones, in view of the higher odds of ESBL-PE acquisition associated with this class.

The study design of this meta-analysis included only prospective cohort studies with robust microbiologic assessments of ESBL-PE carriage. The strength of excluding ESBL-PE carriers pre-travel is that any ESBL-PE carrier can be shown to have acquired the resistant bacteria during travel, which can in turn may generally be attributed to exposures and behaviours taken during the trip. Additionally, while each study recruited travellers from a single country in North America, Europe or Australia, the aggregate analysis supports a broader generalization of healthy tourists from high income regions of the world travelling internationally to tropical, lower income countries.

However, there are limitations to the analysis. First, antimicrobial effect on ESBL-PE acquisition may be mediated partially through an indication of gastroenteritis, which is both an independent risk factor for ESBL-PE acquisition^9,11^ and also a condition for which fluoroquinolones are commonly recommended for international travellers^37^. That is, there is a question about the direction of association between fluroquinolone use and carriage of ESBL-PE. While an analysis accounting for antimicrobial indication was not possible in this review, several prospective studies have adjusted for diarrhea or gastroenteritis as a confounder^9,11,18,19,21^. In these studies, a strong and significant effect of antimicrobial on increased rates of ESBL-PE acquisition during travel persisted. Similarly, many studies documented the presence or absence of proven confounders in individual participants, including age, sex, length of travel, and travel destination which are potentially related to both rates of antimicrobial use, and risk of ESBL-PE acquisition. However, however individual studies including the single largest study included in this review^9^ were able to account for these variables in the analysis, and found a persistent effect on ESBL-PE acquisition related to antimicrobial exposure during travel.

Additionally, the designs of the included studies present potential limitations. For instance, losses to follow-up after travel, while not large where provided, were not reported in 4 studies. Selection bias may occur if those lost had different rates of antimicrobial use and rates of ESBL-PE acquisition; however, it seems unlikely the magnitude of this effect would be large enough to substantially change the observed odds ratio. Moreover, participants in all studies were non-random volunteers from higher income countries, in most cases recruited from pre-travel or vaccination clinics, which may limit generalizability of the results to travellers not attending such clinics or originating from lower income countries. Meanwhile, as antimicrobial exposure was self-reported after travel, and data on antimicrobial class was limited to 11 studies, which might underestimate the effect or precision of our assessment of specific classes during travel on ESBL-PE acquisition.

## CONCLUSION

The odds of acquiring ESBL-PE are substantially increased when antimicrobials are consumed during travel. This risk is shared unevenly among antimicrobial classes, with fluoroquinolones posing a substantial risk compared to others. Incorporation of this analysis in decisions and guidelines addressing whether to use antimicrobials during travel will allow for a better realization of the true risks versus benefits. We call for further study into the mechanisms by which antimicrobials, and in particular fluoroquinolones, contribute to AMR, including indirect mechanisms mediated by disruption of the gut microbiome. This may identify fruitful protective factors which may ameliorate the effect of antimicrobials on AMR, which are needed. In the meantime, it seems prudent for travel health practitioners to emphasize limiting the consumption of antimicrobials for traveller’s diarrhea to treatment of severe cases only.

## Data Availability

There are no external datasets available for this article. Systematic review and meta-analysis of previously published works are included, only.

## Acknowledgements

We would like to offer special thanks to corresponding authors of studies who responded to our requests for further data, which made an invaluable contribution to the precision of our analysis: Drs. Christoph Lübbert, Sunita Paltansing, Thongpan Leangapichart, John Penders, Martin Angelin, Esther Künzli, Karin Ellen Veldkamp, and Marguerite Bruijning.

## BOX 1.

**DETAILED SEARCH STRATEGY USED IN PUBMED**

PubMed

(Enterobacteriaceae[MeSH Major Topic] OR (Enterobacteriaceae*[tiab] OR Gram negative bacteri*[tiab] OR E. coli [tiab]OR Escherischia*[tiab] OR Klebsiella*[tiab] OR Salmonella[tiab] OR Proteus[tiab] OR Enterobacter[tiab] OR Shigella[tiab] OR Yersinia[tiab] OR gut flora[tiab])) AND (Drug Resistance, Bacterial[MeSH Major Topic] OR (Drug resistan*[tiab] OR Extended-spectrum beta-lactamase[tiab] OR ESBL[tiab] OR Amp C[tiab] OR Carbapenemase*[tiab] OR CPE[tiab] OR CRE[tiab] OR Cephalosporinase[tiab] OR Penicillinase[tiab] OR beta-lactamase[tiab] OR CTX-M[tiab] OR ((Cephalosporin[tiab] OR Cefepime[tiab] OR ceftriaxone[tiab] OR Cefotaxime[tiab] OR Ceftazidime[tiab] OR antibiotic*[tiab] OR antimicrob*[tiab]) AND (sensitivit*[tiab] OR susceptibil*[tiab] OR resistan*[tiab])))) AND (Travel[MeSH Major Topic] OR (Travel*[tiab] OR International[tiab] OR Trip[tiab] OR Voyage[tiab] OR Air Transport[tiab] OR Post-Travel[tiab] OR Foreign[tiab] OR Touris*[tiab] OR Aviation[tiab] OR Airport[tiab]))

